# Whole blood-based measurement of SARS-CoV-2-specific T cell responses reveals asymptomatic infection and vaccine efficacy in healthy subjects and patients with solid organ cancers

**DOI:** 10.1101/2021.06.02.21258218

**Authors:** Martin J. Scurr, Wioleta M. Zelek, George Lippiatt, Michelle Somerville, Stephanie E. A. Burnell, Lorenzo Capitani, Kate Davies, Helen Lawton, Thomas Tozer, Tara Rees, Kerry Roberts, Mererid Evans, Amanda Jackson, Charlotte Young, Lucy Fairclough, Mark Wills, Andrew D. Westwell, B. Paul Morgan, Awen Gallimore, Andrew Godkin

**Author notes:** Corresponding Authors*: Prof Andrew Godkin, Division of Infection and Immunity, Henry Wellcome Building, Health Park, Cardiff, CF14 4XN. **Authorship note**: Aw.G and An.G contributed equally to this work (Awen Gallimore, Andrew Godkin).

## Abstract

Accurate assessment of SARS-CoV-2 immunity in the population is critical to evaluating vaccine efficacy and devising public health policies. Whilst the exact nature of effective immunity remains incompletely defined, SARS-CoV-2-specific T cell responses are a critical feature of the immune response that will likely form a key correlate of protection against COVID-19. Here, we developed and optimised a high-throughput whole blood-based assay to determine the T cell response associated with prior SARS-CoV-2 infection and/or vaccination amongst 156 healthy donors and 67 cancer patients. Following overnight *in vitro* stimulation with SARS-CoV-2-specific peptides, blood plasma samples were harvested and analysed for T_H_1-type effector cytokines (IFN-γ and IL-2). Amongst healthy donors, highly significant differential IFN-γ^+^/IL-2^+^ SARS-CoV-2-specific T cell responses were seen amongst vaccinated or previously infected COVID-19-positive individuals in comparison to unknown/naïve individuals (P < 0.0001). IL-2 production from T cells in response to SARS-CoV-2 derived antigens was a highly predictive diagnostic assay (P < 0.0001; 96.0% sensitivity, 93.9% specificity); measurement of IFN-γ^+^ SARS-CoV-2 specific T cell responses was equally effective at identifying asymptomatic (antibody and T cell positive) participants. A single dose of COVID-19 vaccine induced IFN-γ and/or IL-2 SARS-CoV-2-specific T cell responses in 28/29 (96.6%) of healthy donors, reducing significantly to 27/56 (48.2%) when measured in cancer patients (P = 0.0003). Overall, this cost-effective standardisable test ensures accurate and comparable assessments of SARS-CoV-2-specific T cell responses amenable to widespread population immunity testing.

## Background

As the COVID-19 pandemic continues, there is an increasing focus on understanding how adaptive immune responses generated from severe acute respiratory syndrome coronavirus 2 (SARS-CoV-2) infection and/or vaccination provide protection from future infection. Although the exact determinants of effective immunity from re-infection with SARS-CoV-2 remain to be deciphered, multiple recent studies have revealed that virus-specific T cell responses develop in nearly all individuals with confirmed SARS-CoV-2 infection (1-4), with responses persisting for at least six months post-infection (5). Traditional means of assessing viral antigen-specific T cell responses utilise flow cytometry or ELISpot-based readouts; however, neither approach is standardisable across multiple laboratories, cost-effective, or amenable to high-throughput processing, thus precluding their use for larger scale population immunity screen. In addition, current commercially available immunoassays that detect cellular immune responses to SARS-CoV-2 solely measure IFN-γ released by antigen-specific T cells (6), even though other T_H_1-type cytokines may be better indicators of anti-viral response (5). To overcome these limitations, existing whole blood-based *in vitro* immunodiagnostics, such as those used for measuring Mycobacterium tuberculosis-specific T cell responses (7), can be adapted to measure virus-specific T cell responses in a high-throughput, standardisable manner. Specifically, this T cell immunoassay measures cytokines in the plasma released by antigen-specific T cells following stimulation with specific peptides spanning antigenic regions of the pathogen. This approach is gaining recognition as a potentially powerful diagnostic tool for managing the COVID-19 pandemic (8-10).

Monitoring immunological responses to SARS-CoV-2 is of particular importance amongst the elderly and immunosuppressed, given the significantly higher mortality rates observed in these groups (11). Recent studies have associated higher rates of COVID-19 morbidity and mortality with sub-optimal adaptive immune responses (12). The incidence of cancer is also increased in the elderly where a declining adaptive immune system, as well as age-associated inflammation, are factors in disease progression. Given that influenza vaccines induce weaker immune responses in the elderly and in cancer patients (13, 14), measuring adaptive immune responses to SARS-CoV-2 in vaccinated individuals belonging to these groups is important. Indeed, early indications suggest that cancer patients, in particular those on active treatments such as chemotherapy, were significantly less likely to mount antibody and T cell responses to the Pfizer-BioNTech SARS-CoV-2 mRNA vaccine, (15).

Here, we adapted and optimised a widely utilised *in vitro* whole blood stimulation assay to determine the presence of SARS-CoV-2-specific T_H_1-type (IFN-γ / IL-2) cellular immune responses in healthy donors, to assess T cell responses generated from prior infection, whether the participant was symptomatic or not, and as a readout of vaccine efficacy amongst healthy donors and cancer patients. We demonstrate high sensitivity and specificity of this assay to identify or exclude prior SARS-CoV-2 infection and/or successful COVID-19 vaccination. Going forward, it is imperative to utilise such tests to understand the precise contribution of T cell responses with regards to long-term immunity to SARS-CoV-2 infection, in particular amongst immunocompromised individuals.

## Methods

### Study cohorts

Participants were recruited to this research project between February-April 2021. A healthy donor cohort (n = 156) comprised university staff and students attending Cardiff University’s COVID-19 Screening Service or members of the public attending a Cardiff-based GP practice. All participants were otherwise healthy and did not report taking any current immunosuppressive treatment. In addition, patients with a range of solid-organ cancers (n = 67) were recruited from Velindre Cancer Centre prior to receiving their first COVID-19 vaccine (see Table 1 for basic characteristics). All participants were stratified based on self-reported and/or laboratory evidence of a prior SARS-CoV-2 infection. Participants reporting no prior positive test were defined as ‘unknown/naïve’. To measure immunological responses generated to COVID-19 vaccination, baseline blood samples were taken immediately preceding the first dose; an additional post-vaccination blood sample was taken 3-6 weeks later. Only participants donating a blood sample both pre- and post-vaccination were included in the analyses. All vaccinated participants received either Pfizer (BNT162b2) mRNA vaccine or AstraZeneca (ChAdOx1 nCoV-19) vaccine.

**Table 1.** Basic participant characteristics

|  | **Cancer patients (n=68)** | **Healthy donors (n=156)** |
| --- | --- | --- |
| Mean age, years (range) | 52.6 (28-79) | 35.6 (18-81) |
| **Sex** |  |  |
| Male | 25/68 (36.8%) | 48/156 (30.8%) |
| Female | 43/68 (63.3%) | 108/156 (69.2%) |

This study received ethical approval from the Wales Cancer Bank (WCB No. 21/004), the Newcastle & North Tyneside 2 Research Ethics Committee (IRAS ID: 294246) and Cardiff University School of Medicine Research Ethics Committee (SREC reference: SMREC 21/01). All participants gave written, informed consent prior to inclusion.

### Peptides

All SARS-CoV-2 peptides were dissolved according to manufacturer’s instructions (Miltenyi Biotec). A single SARS-CoV-2-specific S-/NP-/M-combined peptide pool was created, comprising mainly 15-mer sequences with 11-amino acid overlap, covering the entire spike (S1 and S2) protein (S), nucleocapsid phosphoprotein (NP) and membrane glycoprotein (M). All peptides were used at a final concentration of 0.5 μg/ml.

### Stimulation

A single 6ml or 10ml sodium heparin vacutainer (BD) tube of venous blood was collected from each participant and processed in the laboratory within 12 hours of blood draw. Whole blood samples were aliquoted into microcentrifuge tubes (Thermo Scientific) containing pre-aliquoted peptides, alongside additional tubes containing phytohaemagglutinin (Sigma) (positive control) or nothing (negative control). Samples were incubated at 37°C for 20-24 hours. Tubes were then centrifuged at 3000g for 2 minutes before harvesting plasma from the top of each blood sample. Plasma samples were stored at -20°C for up to one month prior to running cytokine detection assays.

### Detection of anti-SARS-CoV-2 RBD IgG Antibodies

An in-house direct ELISA was developed as previously described (16-19), with some modifications. Maxisorp (Nunc, Loughborough, UK) 96-well plates were coated with RBD protein (recombinantly generated in a mammalian expression system, in house) at 2 μg/ml in bicarbonate buffer, pH 9.6 at 4°C overnight; wells were blocked for 1 hour at room temperature with 3% w/v non-fat dried milk powder (Sigma Aldrich, # 70166-500G) in phosphate-buffered saline containing 0.1% Tween 20 (PBS-T), washed in PBS-T. Dilutions of patient sera (1 in 50 in 1% Milk PBS-T), were added in duplicate to wells coated with RBD protein and incubated for 2 hours at room temperature. Wells were washed three times with PBS-T then incubated (1 hour, room temperature) with secondary antibody (donkey anti-human IgG F(ab’)_2_-horseradish peroxidase (HRP); #709-036-149, Jackson ImmunoResearch, Ely, UK) for 1 hour at room temperature. After washing (x3), plates were developed using O-phenylenediamine dihydrochloride (OPD, SIGMAFASTTM; Sigma-Aldrich, # P9187-50SET) and the optical density (OD) measured at 492 nm. Assay validation, including intra-/inter-assay CVs for this assay have been previously described (17, 19).

### Cytokine Detection

IFN-γ was measured using the IFN-γ ELISA MAX Deluxe kit (BioLegend) and performed according to manufacturer’s instructions. Microplates were read at 450nm immediately following the addition of stop solution (2N H_2_SO_4_). IFN-γ was quantified by extrapolating from the standard curve using Graphpad Prism. Values below the lower limit of detection of the assay were recorded as 7.81 pg/ml. IL-2 was measured using a custom Bio-Plex Pro Human Cytokine Set (Bio-Rad) and performed according to manufacturer’s instructions. The mean fluorescent intensity of the cytokine beads was measured on a Bio-Plex 200 (Bio-Rad). Cytokine concentration was calculated from control curves of standards provided in the kit. Values below the lower limit of detection of the assay were recorded as 6.28 pg/ml.

### Statistics

GraphPad Prism Version 9 was used for all statistical analyses of datasets. Dataset normality was tested using the Shapiro-Wilk test. Significance was determined using either Kruskal-Wallis tests with corrections for multiple comparisons made using the Dunn test, or Mann-Whitney tests, as indicated in the figure legends. Correlation analyses were performed using linear regression. All tests were performed two-sided with a nominal significance threshold of P < 0.05.

## Results

Prior analysis of the SARS-CoV-2-specific T cell cytokine profile, measured in supernatants of *ex vivo* peptide-stimulated ELISpot cultures or whole blood assays, revealed that T_H_1-type IFN-γ^+^ and IL-2^+^ responses dominate effective, functional SARS-CoV-2-specific T cell responses (5, 10). The magnitude of T_H_1-type responses generated to the SARS-CoV-2 S-/NP-/M-combined peptide pool was assessed amongst healthy donor participants who had either received a prior vaccination (n = 52), a confirmed prior infection (n = 15) (PCR positive swab from nasopharynx or saliva sample and/or a strong history of infection with positive measured antibody responses to SARS-CoV-2) or no known history of infection (n = 88). Highly significant differential IFN-γ-positive and IL-2-positive SARS-CoV-2-specific T cell responses were seen amongst vaccinated or previously infected (non-vaccinated) COVID-19-positive individuals in comparison to unknown/naïve individuals (Figure 1A; P < 0.0001 and Figure 1B; P < 0.0001, respectively). There was no significant difference in the magnitude of IFN-γ or IL-2 production between vaccinated and infected participants (Figure 1A and 1B, respectively). Amongst all vaccinated, infected or unknown/naïve participants, there were highly significant correlations between IFN-γ and IL-2 production, despite a propensity for increased (>20 pg/ml) IFN-γ^+^ SARS-CoV-2-specific T cell responses without a measurable IL-2 response amongst unknown/naïve donors (Figure 1C).

**Figure 1.**
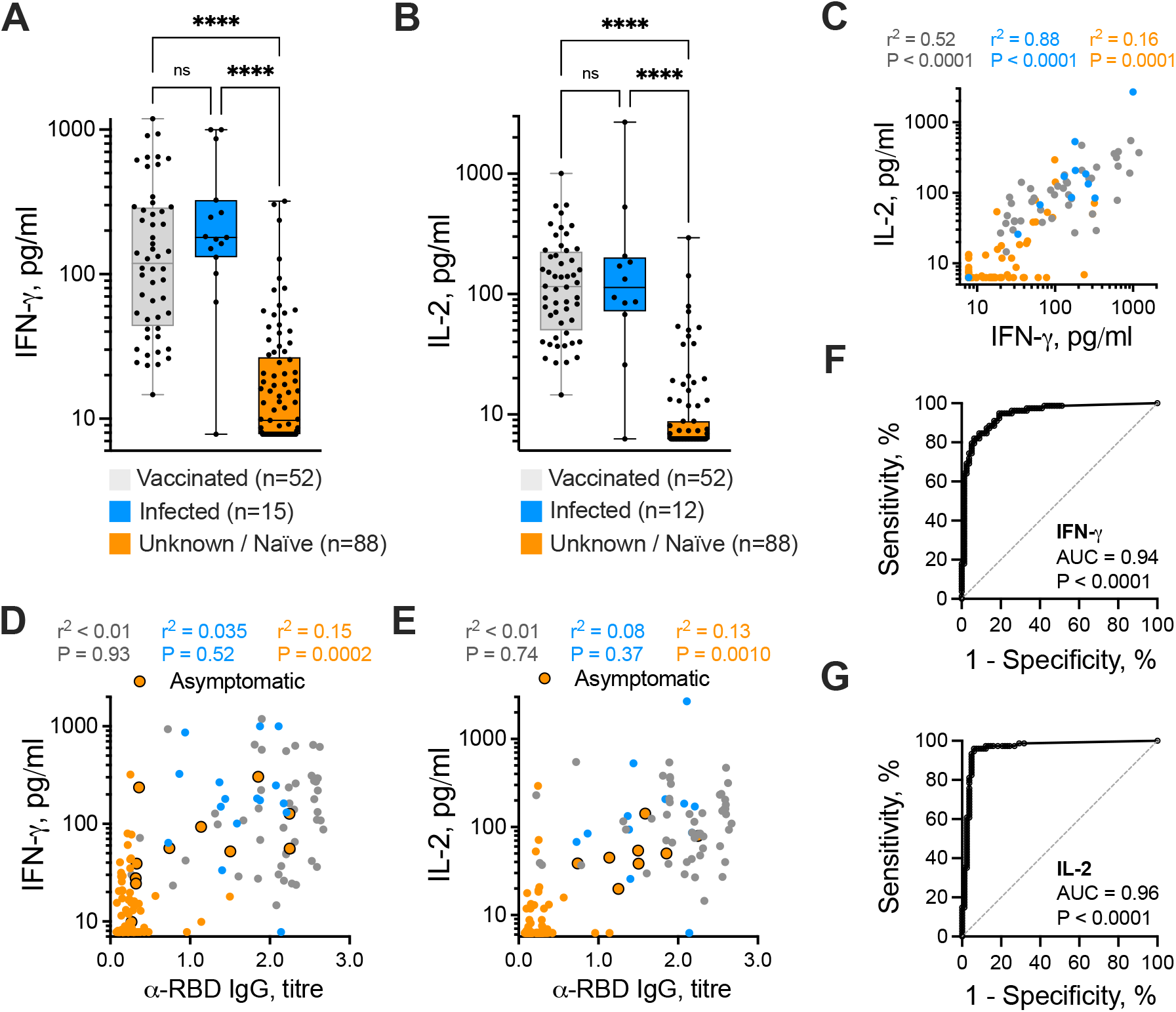
SARS-CoV-2-specific T cell response identifies prior asymptomatic infection. IFN- (A) and IL-2 (B) release in response to the SARS-CoV-2 S-/NP-/M-combined peptide pool was measured in 156 evaluable participants, subdivided into those with prior COVID-19 vaccination (grey, n = 52), prior COVID-19-positive PCR test result (blue, n = 12-15), or those with no prior positive COVID-19 test, termed ‘unknown/naïve’ (orange, n = 88). Statistical analyses indicate the results of a Kruskal-Wallis test with Dunn’s correction for multiple comparisons (**** P < 0.0001). (C) The SARS-CoV-2-specific IL-2^+^ T cell response was correlated with the IFN-γ^+^ response, subdivided by participant status. The anti-SARS-CoV-2 RBD IgG antibody titre was correlated with the magnitude of IFN-γ^+^ T cell response (D) and IL-2^+^ T cell response (E), with identified asymptomatic participants highlighted. Results of regression analyses are indicated. Sensitivity and specificity readouts for IFN-γ (F) and IL-2 (G) were defined by receiver operating characteristic curves. Area under the curve (AUC) and associated P value are indicated.

Next, we correlated participant T cell responses with anti-SARS-CoV-2 RBD IgG antibodies. There was a strong concordance, albeit not significant correlation, between the presence of both cellular and humoral immune responses in all vaccinated and previously infected participants: only 1/15 (6.7%) previously infected participant had a positive antibody response (>0.27) without a measurable IFN-γ ^+^ or IL-2^+^ T cell response. All 52 vaccinated participants demonstrated a measurable IFN-γ^+^or IL-2^+^ T cell response, although two were antibody negative (<0.27) (Figures 1D and 1E). To investigate whether the increased T cell responses observed amongst naïve/unknown participants with no history of confirmed infection was indicative of asymptomatic SARS-CoV-2 infection, we further compared the magnitude of IFN-γ ^+^or IL-2^+^ T cell responses with evidence of antibody seroconversion; a significant correlation between T cell response and the magnitude of anti-SARS-CoV-2 RBD IgG antibodies was noted for unknown/naïve participants (IFN-γ : P = 0.0002, Figure 1D; IL-2: P = 0.0010, Figure 1E). Ten such participants were identified as having both a positive antibody response (>0.27) and raised (above median) IFN-γ ^+^T cell response, as indicated (Figure 1D), and eight participants also had a positive antibody and increased IL-2^+^ T cell response (Figure 1E), consistent with the adaptive immune responses generated in the majority of vaccinated or previously infected participants. Given these participants self-reported no prior confirmed COVID-19 test or symptoms associated with COVID-19 during the pandemic, the presence of both a SARS-CoV-2-specific T cell and antibody response is highly indicative of prior asymptomatic infection.

When assessing the utility of measuring SARS-CoV-2-specific T cell responses to identify those who have received prior COVID-19 vaccination and/or been previously infected, Youden’s index revealed an optimal cut-off value of >23.55 pg/ml IFN-γ, achieving a sensitivity of 93.59% (95% CI 85.9-97.2%) and specificity of 80.8% (95% CI 70.7-88.0%) (AUC = 0.94 (95% CI 0.91-0.98); P < 0.0001; Figure 1F).

Measuring IL-2^+^ T cell response as the readout adjusted the sensitivity and specificity to 96.0% (95% CI 88.8-98.9%) and 93.9% (95% CI 86.5-97.4%), respectively, at an optimal cut-off value of >20.17 pg/ml IL-2 (AUC = 0.96 (95% CI 0.93-1.00); P < 0.0001; Figure 1G).

Next, we further evaluated eleven non-vaccinated COVID-19-positive convalescent participants reporting mild to moderate severity of associated symptoms, and ten non-vaccinated participants reporting no prior associated symptoms and for whom both IFN-γ and IL-2 data was available (Figure 2A), to determine the functionality of SARS-CoV-2-specific T cells induced by prior asymptomatic infection. The magnitude of IFN-γ production was reduced in asymptomatic participants (P = 0.061, Figure 2A and 2B); furthermore, a significant reduction in IL-2 production was noted (P = 0.0011, Figure 2A and 2C). When assessing all T_H_1 responses by participant infection and symptom status and using optimal cut-offs defined above to identify positive responses, dual-producing IFN-γ^+^/IL-2^+^ SARS-CoV-2-specific T cell responses were present in 10/11 (90.9%) of symptomatic participants, reducing to 7/10 (70.0%) amongst asymptomatic participants (Figure 2D). Given the diminished nature of the SARS-CoV-2-specific T_H_1-type cytokine production in asymptomatic donors, both IFN-γ and IL-2 are equally useful readouts when assessing for the presence of T cell responsiveness.

**Figure 2.**
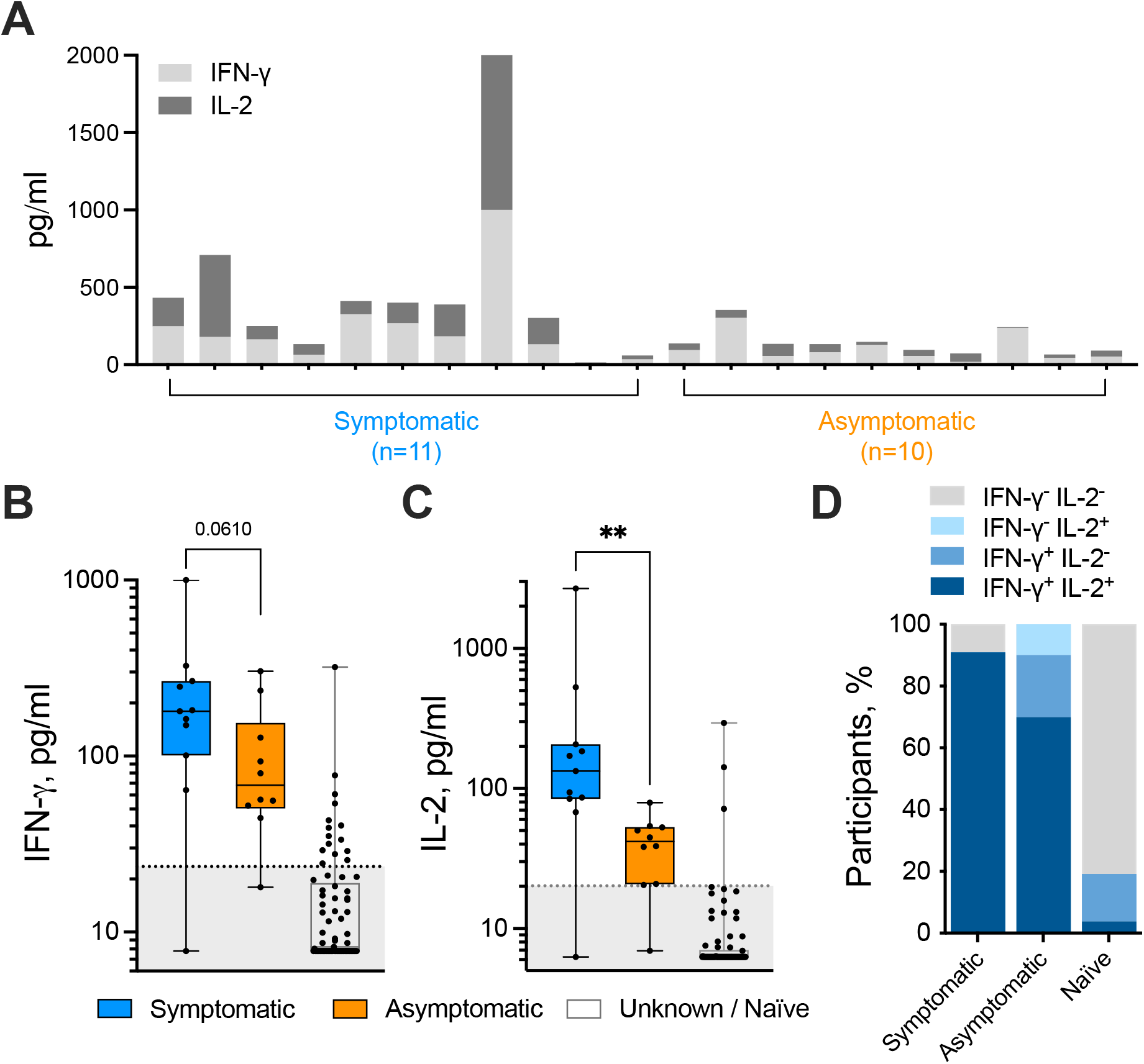
Asymptomatic participants exhibit reduced SARS-CoV-2-specific T cell functionality. (A) IFN-γ and IL-2 T cell responses amongst eleven symptomatic and 10 asymptomatic SARS-CoV-2-infected donors is shown for each individual. IFN-γ (B) and IL-2 (C) release by T cells in response to the SARS-CoV-2 S-/NP-/M-combined peptide pool was measured in the symptomatic, asymptomatic and naïve donors (n = 76). P values resulting from Mann Whitney tests are shown (** P < 0.01). The proportion of symptomatic, asymptomatic and naïve participants mounting dual IFN-γ^+^/IL-2^+^, single IFN-γ or IL-2^+^ or no measurable T cell response is shown (D).

SARS-CoV-2-specific T cell responses amongst unknown / naïve participants, i.e. those with no measurable anti-SARS-CoV-2 RBD IgG antibodies and no prior confirmed history of infection, were rare, with 3/76 (3.8%) displaying a dual IFN-γ^+^/IL-2^+^ response and 12/76 (15.8%) displaying an IFN-γ^+^/IL-2^-^ response. This could be indicative of pre-existing, cross-reactive T cells, or a terminally differentiated effector T cell response. However, further studies are required to ascertain whether those participants with raised anti-SARS-CoV-2-specific T_H_1 responses without seroconversion have been infected by SARS-CoV-2. Interestingly, this discordant immune response has also been reported when monitoring intra-familial exposure to the virus (20).

Finally, we tracked T cell and antibody responses pre- and post-single dose SARS-CoV-2 vaccination amongst a cohort of healthy controls and a cohort of cancer patients with solid tumours. Robust priming of SARS-CoV-2-specific T cells was observed amongst healthy controls, with 28/29 (96.6%) mounting an IFN-γ ^+^ response >23.55 pg/ml (Figure 3A) and 20/22 (90.9%) mounting IL-2^+^ responses >20.17 pg/ml (Figure 3B). In contrast, only 27/56 (48.2%) and 18/31 (58.1%) of cancer patients mounted an IFN-γ (Figure 3A) or IL-2 (Figure 3B) response following single dose SARS-CoV-2 vaccination, respectively. Furthermore, anti-SARS-CoV-2 RBD IgG antibody responses were also compromised in cancer patients though to a lesser degree than the T cell response, with 42/54 (77.8%) of cancer patients vs. 27/28 (96.4%) of healthy controls reaching the threshold of positivity in our antibody test (Figure 3C). This may reflect a limitation of the whole blood-based SARS-CoV-2 T cell assay in that insufficient numbers of lymphocytes may be present to detect such a response; this may be a particular problem in cancer given that lymphopenia is a common occurrence. However, such a situation is also likely to hamper the induction of long-term T cell memory formation.

**Figure 3.**
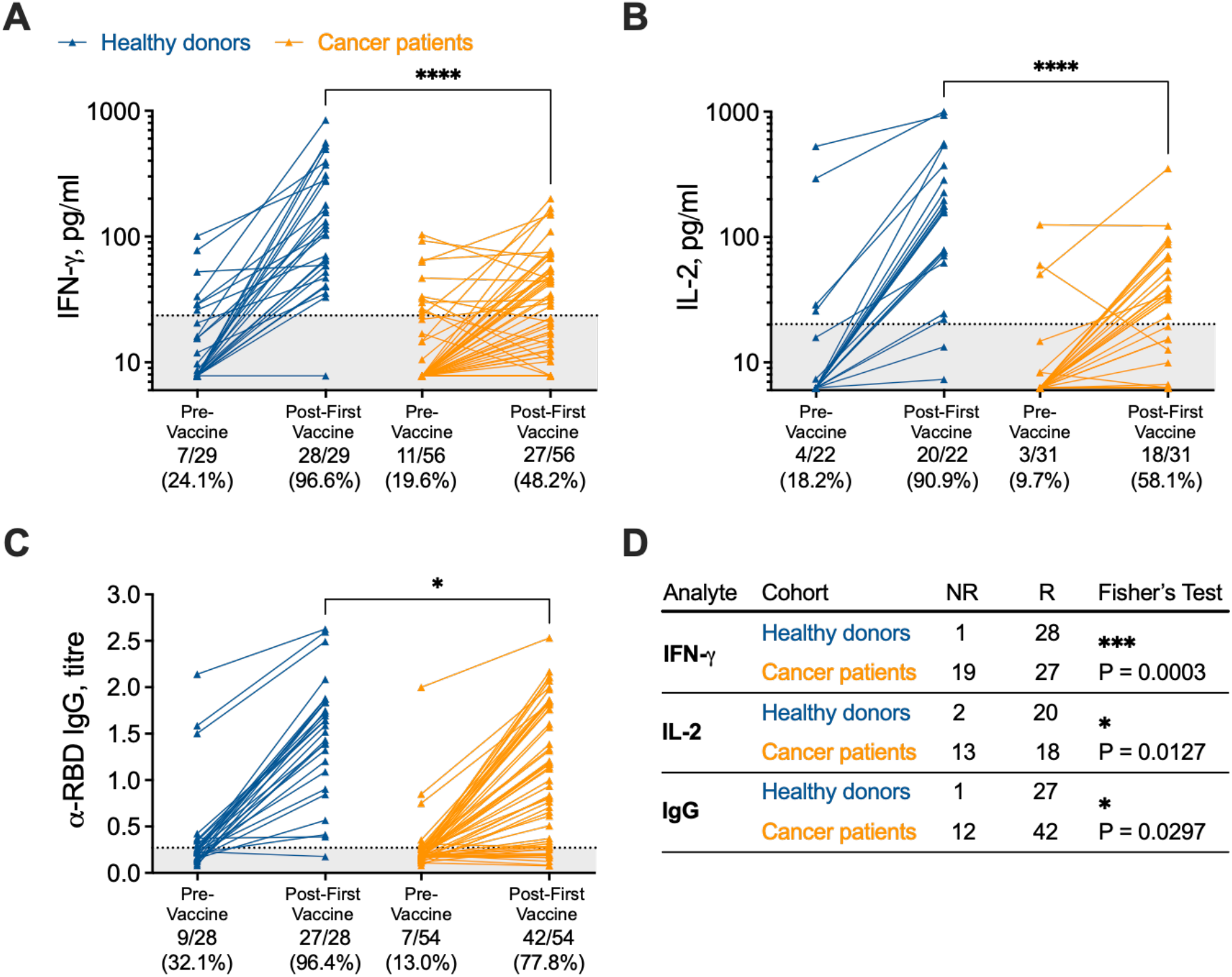
Utilising SARS-CoV-2-specific T cell response measurements as a readout for COVID-19 vaccine efficacy amongst healthy donor and cancer patient cohorts. IFN-γ^+^(A) and IL-2^+^ (B) SARS-CoV-2-specific T cell responses and anti-SARS-CoV-2 RBD IgG levels (C) were measured before and 3-6 weeks after a single COVID-19 vaccination in healthy donors and cancer patients. Two-tailed P values resulting from Mann Whitney tests are shown. (D) Vaccination response rates (NR = no response, R = response) in terms of positive T cell (IFN-γ, IL-2) or antibody (RBD IgG) response amongst each cohort is shown. P values indicate results from Fisher’s exact test.

Overall, these data highlight the power of measuring SARS-CoV-2-specific T cell responses as a means for identifying prior COVID-19 infection, vaccination efficacy and/or potential future immunity from re-infection status.

## Discussion

This study demonstrates the utility of a high-throughput, standardisable T cell immunoassay to accurately detect SARS-CoV-2-specific T cell responses. In order to control future outbreaks and identify at-risk individuals, the exact constituents of effective COVID-19 immunity at a population level must be understood. When used alongside measurements of virus-specific antibodies, T cell response readouts represent a powerful, additional measure of potential immunity from COVID-19, with a higher degree of confidence than either measurement on their own, in particular given the concern on the longevity of measurable antibody responses (21-23). In addition, the FDA’s decision to issue emergency use authorisation for a SARS-CoV-2 T cell test highlights the growing acceptance and usefulness of T cell testing for the clinical management of certain patient groups (24).

Here, we show that measuring plasma T_H_1-type effector cytokines from SARS-CoV-2 peptide-stimulated whole blood can accurately detect the presence of a cellular immune response to SARS-CoV-2, distinguishing those who have received prior vaccination and/or infection from uninfected healthy donors with a high degree of sensitivity and, in particular for IL-2, specificity. These results are consistent with comprehensive analyses of the cytokine profile released by SARS-CoV-2-specific T cells measured in whole blood-based assays or ELISpot / cell culture supernatants, which showed that IL-2 and IFN-γ are the dominant cytokines (5, 10, 25). However, the relevance of an IFN-γ-positive, IL-2-negative SARS-CoV-2-specific T cell response in uninfected donors with respect to long-term immunity requires further investigation. The peptides used in our immunoassay predominantly stimulate CD4^+^ T cell responses, and cover all major immunodominant regions of the virus, including those in the spike, membrane and nucleocapsid proteins, as recently defined (26). T_H_1-type responses to these immunogenic regions were detected in the vast majority of convalescent SARS-CoV-2 infected and/or vaccinated individuals, in keeping with prior findings (26, 27), although convalescent asymptomatic donors and a minority of naïve individuals demonstrate low-level reactivity. Prior studies have revealed the functionality and magnitude of adaptive immune responses to SARS-CoV-2 were significantly lower in mild cases of COVID-19 in comparison to severe (5), potentially the result of lower viral loads (27, 28). In accordance with this, our study revealed that production of IFN-γ and IL-2 was lower in COVID-19-convalescent asymptomatic participants. However, a recent longitudinal analysis of asymptomatic SARS-CoV-2 infection identified an increased, highly functional IFN-γ and IL-2 response within 3 months of infection that declined faster than in symptomatic individuals (10). Differences in post-infection sampling times likely account for the discrepancy in associations between SARS-CoV-2-specific T cell responses and COVID-19 symptom severity, given that the majority of our participants were infected ∼6 months prior to blood sampling. However, it is encouraging to note that T cell responses were still present and functional over this timeframe in nearly all SARS-CoV-2-infected convalescent and/or vaccinated participants, even in those up to 12 months post-infection, corresponding well with prior studies describing the longevity of anti-SARS-CoV-2 T cell immunity (5). Whether these responses provide immune protection, especially to new emerging mutant variants of the SARS-CoV-2 virus, warrants further investigation in larger prospective cohorts. In addition, further downstream analyses incorporating other coronaviruses and additional antigenic regions are necessary to ascertain whether cross-reactive T cell responses play a role in SARS-CoV-2 immunity.

Future studies are on-going to evaluate the durability of these measured T cell responses in our participants, comparing healthy subjects with immunocompromised patients, such as those with cancer. After the first dose of vaccine in cancer patients, a significantly weaker induction of cellular and, to a lesser extent, humoral responses was found, corroborating similar observations in other studies (15). These data provide further support to recent calls for cancer patients to be prioritised for booster vaccines and longer-term immunological monitoring.

In summary, we describe an immunoassay that accurately and rapidly identifies the presence of SARS-CoV-2-specific T cell responses, both helping to elucidate the adaptive immune status of previously infected and/or vaccinated individuals, and diagnosing previously unsuspected past infection. Incorporating qualitative T cell response data in population immunity studies, or individualised certifications of immunity, could have far reaching implications for government policy on lockdown restrictions, and more effectively assess vaccine efficacy in communities, highlighting the potential requirement for repeat vaccinations where immunity wanes.

## Supporting information

STROBE Statement

## Data Availability

The data that support the findings of this study are available on request from the corresponding author.

## Acknowledgements

Biosamples from cancer patients were sourced through the Wales Cancer Bank (DOI: http://doi.org/10.5334/ojb.46) which is funded by Health and Care Research Wales. Other investigators may have received specimens from the same subjects.

Our thanks are extended to all participants in this study.

